# Joint Longitudinal-Survival Modelling of Patient-Reported Gastrointestinal Symptom Trajectories and Treatment Discontinuation in Irritable Bowel Syndrome: A Prospective Cohort Study from the Canadian Gut Project

**DOI:** 10.64898/2026.03.16.26348556

**Authors:** Emily Thornton, Jonathan Kellerman

## Abstract

**Background:** Irritable bowel syndrome (IBS) is characterized by heterogeneous symptom trajectories and high treatment discontinuation rates. Traditional analyses examine longitudinal outcomes and time-to-event endpoints separately, potentially missing informative dropout and the association between symptom dynamics and treatment persistence.

**Objective:** To jointly model patient-reported IBS symptom trajectories and time-to-treatment discontinuation using shared random effects, characterizing the association between individual symptom dynamics and treatment persistence in a large Canadian prospective cohort.

**Methods:** We analyzed 2,847 adults with Rome IV diagnosed IBS enrolled in the Canadian Gut Project (2018 to 2024) across 14 gastroenterology centres in Alberta, British Columbia, and Ontario. The longitudinal submodel used linear mixed-effects regression for the IBS Severity Scoring System (IBS-SSS) measured at baseline and months 3, 6, 12, 18, and 24. The survival submodel used a Weibull proportional hazards model for time-to-treatment discontinuation. The joint model linked both processes through shared random effects (random intercept and slope), estimated via maximum likelihood with adaptive Gauss-Hermite quadrature (15 nodes). We conducted sensitivity analyses using Bayesian estimation, alternative association structures (current value, time-dependent slopes), and multiple imputation for intermittent missingness.

**Results:** Mean baseline IBS-SSS was 298.4 (SD 72.1). Over 24 months, 1,042 participants (36.6%) discontinued treatment. The longitudinal submodel revealed a mean IBS-SSS decline of −8.7 points/month (95% CI: −10.2, −7.1) with substantial between-person heterogeneity in both intercepts (STD = 4,218.3) and slopes (STD = 12.4). The association parameter linking the shared random intercept to the hazard of discontinuation was α₁ = 0.0034 (95% CI: 0.0021, 0.0047; p < 0.001), indicating that each 10-point increase in individual-specific baseline severity increased the hazard of discontinuation by 3.5%. The shared slope association parameter was α₂ = −0.187 (95% CI: −0.264, −0.110; p < 0.001), demonstrating that individuals with steeper symptom improvement had lower discontinuation hazards. IBS-D subtype (HR = 1.41; 95% CI: 1.18, 1.69), concurrent anxiety (HR = 1.28; 95% CI: 1.09, 1.50), and social media health information use (HR = 0.82; 95% CI: 0.71, 0.95) were significant predictors in the survival submodel.

**Conclusion:** Joint longitudinal-survival modelling reveals that IBS symptom trajectories and treatment discontinuation are dynamically linked through individual-level latent processes. Higher baseline severity and slower improvement trajectories significantly predict earlier discontinuation. These findings support personalized treatment monitoring approaches that use real-time symptom trajectory data to identify patients at risk of discontinuation.

## Introduction

Irritable bowel syndrome (IBS) affects an estimated 11.2% of the global population and represents one of the most prevalent functional gastrointestinal disorders, imposing substantial burden on patients and healthcare systems [1]. Characterized by recurrent abdominal pain associated with altered bowel habits, IBS presents as a chronic, relapsing condition with markedly heterogeneous symptom profiles across individuals and over time [2]. The Rome IV diagnostic criteria classify IBS into four subtypes—diarrhoea-predominant (IBS-D), constipation-predominant (IBS-C), mixed (IBS-M), and unsubtyped (IBS-U)—yet even within subtypes, patients demonstrate highly variable longitudinal symptom trajectories that complicate treatment planning and outcome prediction [3].

Treatment persistence in IBS remains a critical clinical challenge. Studies consistently report treatment discontinuation rates of 30–50% within two years across pharmacological, dietary, and psychological interventions [4]. This high attrition undermines therapeutic efficacy and clinical trial validity, yet the dynamic relationship between symptom evolution and discontinuation behaviour is poorly understood. Traditional analyses have addressed these endpoints separately: longitudinal models for symptom trajectories and survival models for time-to-discontinuation.

This separated approach ignores two fundamental issues. First, treatment discontinuation may be informative—patients who discontinue may do so precisely because of their symptom trajectory—violating the missing-at-random assumption of standard mixed models [5]. Second, changes in symptom severity over time may influence the hazard of discontinuation in ways that cannot be captured by time-varying covariates alone, as the latent individual-specific trajectory parameters (intercepts and slopes) may be the true drivers of the event process [6].

Joint longitudinal-survival modelling provides a principled statistical framework for simultaneously estimating both processes while accounting for their interdependence through shared latent structures [7]. By linking individual-specific random effects from the longitudinal submodel to the hazard function in the survival submodel, joint models correct for informative dropout, improve efficiency in estimating longitudinal trajectories, and enable dynamic prediction of event probabilities conditional on observed longitudinal data [8]. Developed initially in HIV/AIDS research for modelling CD4 counts and time-to-clinical events, this methodology has since been extended to oncology, cardiovascular medicine, and nephrology, yet its application in functional gastrointestinal disorders remains nascent [9].

The need for nuanced understanding of IBS patient experiences has been underscored by recent social media discourse analyses. Shankar and Yip (2025) demonstrated through sentiment analysis and topic modelling of IBS-related posts on X.com that patient discussions encompass eight major thematic domains—from physical symptoms and dietary management to social support needs and mental health concerns—with sentiment distributions revealing that 18.7% of discourse conveyed negative sentiment reflecting frustration, despair, and struggles with ongoing symptom management [10]. Their finding that comorbid conditions (12.2% of discourse) and mental health dimensions (7.2%) featured prominently in patient narratives supports the hypothesis that both psychosocial factors and symptom dynamics may jointly influence treatment persistence decisions. Understanding how these patient-reported experiences evolve longitudinally and connect to discontinuation behaviour requires analytical methods capable of modelling both processes simultaneously.

In this study, we apply joint longitudinal-survival modelling with shared random effects to a large prospective cohort from the Canadian Gut Project. Our objectives are threefold: (1) to characterize IBS symptom severity trajectories using linear mixed-effects models with individual-specific random intercepts and slopes; (2) to estimate time-to-treatment discontinuation using parametric survival models; and (3) to quantify the association between individual-level symptom dynamics and discontinuation hazards through shared latent processes, while identifying clinical, psychosocial, and behavioural predictors of both outcomes.

## Methods

### Study Design and Setting

This prospective cohort study was conducted within the Canadian Gut Project, a multicentre longitudinal initiative established in 2017 to investigate functional gastrointestinal disorders across Canada [11]. Participants were enrolled from 14 gastroenterology centres across three provinces: Alberta (n = 5 centres), British Columbia (n = 4), and Ontario (n = 5), between January 2018 and December 2021, with follow-up through December 2024. The study received ethics approval from the Conjoint Health Research Ethics Board at the University of Calgary (REB18-0472) and equivalent institutional review boards at participating sites. All participants provided written informed consent. The study is reported following the Strengthening the Reporting of Observational Studies in Epidemiology (STROBE) guidelines [12].

### Participants

Adults aged 18–75 years with a confirmed diagnosis of IBS according to Rome IV criteria were eligible for inclusion. Exclusion criteria were: (1) diagnosis of inflammatory bowel disease, coeliac disease, or microscopic colitis; (2) history of gastrointestinal malignancy or major abdominal surgery within the preceding 12 months; (3) current pregnancy; (4) inability to complete English- or French-language patient-reported outcome measures; and (5) concurrent participation in an interventional clinical trial. From 3,412 screened individuals, 2,847 met all criteria and were enrolled.

## Measurements

### Primary longitudinal outcome

The IBS Severity Scoring System (IBS-SSS) was administered at baseline and at 3, 6, 12, 18, and 24 months [13]. The IBS-SSS generates a composite score (0– 500) from five dimensions: abdominal pain severity, pain frequency, bloating severity, bowel habit dissatisfaction, and quality-of-life interference. Scores are categorized as remission (<75), mild (75–175), moderate (175–300), and severe (>300).

### Primary survival outcome

Time-to-treatment discontinuation was defined as the interval from enrolment to the first occurrence of voluntary cessation of the prescribed treatment regimen for ≥30 consecutive days, assessed through electronic medical records and patient self-report at each study visit. Participants who did not discontinue treatment were censored at their last study visit or at 24 months, whichever occurred first.

### Covariates

Baseline covariates included: demographics (age, sex, province of residence, household income quartile, education level); clinical characteristics (IBS subtype, symptom duration, number of prior treatments, body mass index, concurrent medications); psychological measures (Hospital Anxiety and Depression Scale [HADS] anxiety and depression subscales); diet (adherence to low-FODMAP diet, assessed by a validated dietary screener); physical activity (International Physical Activity Questionnaire short form); and health information behaviour (self-reported use of social media for IBS-related health information, frequency of online health community participation).

## Statistical Analysis

### Joint modelling framework

We specified a joint model linking a linear mixed-effects (LME) longitudinal submodel for IBS-SSS and a Weibull proportional hazards survival submodel for time-to-treatment discontinuation through shared random effects [7,14].

### Longitudinal submodel

The LME model for subject i at time t□□was specified as: Y□(t□□)=β□ + β□t□ + β□X□ + β□(t□ × X□) + b□ + b□t□ + ε□(t□), where β□ is the fixed intercept, β□ is the fixed slope (average rate of change), X□ is a vector of baseline covariates with coefficients β□ and time interactions β□, and (b□, b□)□ ∼ N(0, D) are subject-specific random intercept and slope with unstructured covariance matrix D. The residual ε□(t□) ∼ N(0, σ^2^) captures within-subject measurement error.

### Survival submodel

The hazard function for subject i was: h□(t) = h□(t) × exp(γ□W□ + α□b□ + α□b□), where h□(t) = κλ(λt)□^1^ is the Weibu□ baseline hazard with shape κ and scale λ, W□ is a vector of baseline covariates with log-hazard ratios γ, and α□ and α□ are the association parameters linking the shared random intercept and slope to the hazard of discontinuation.

### Estimation

The joint likelihood was maximized using the expectation-maximization (EM) algorithm with adaptive Gauss-Hermite quadrature (15 nodes) for numerical integration over the random effects distribution [15]. Standard errors were obtained from the observed information matrix. All analyses were performed in R version 4.3.2 using the JMbayes2 package [16]. We also fitted separate (non-joint) longitudinal and survival models to assess the impact of ignoring the joint structure.

### Alternative association structures

In sensitivity analyses, we examined three additional association structures: (1) current value parameterization, where the hazard at time t depends on the current fitted value of the longitudinal outcome m□(t); (2) current value plus time-dependent slope, incorporating both m□(t) and dm□(t)/dt; and (3) cumulative effects, where the hazard depends on the area under the subject-specific trajectory ∫m□(s)ds. Model comparison used the deviance information criterion (DIC) and Watanabe-Akaike information criterion (WAIC) [17].

### Missing data

Intermittent missingness (i.e., observed data before and after a missed visit) was handled using multiple imputation by chained equations (MICE) with 20 imputed datasets, pooling results using Rubin’s rules [18]. The primary analysis used the full-information maximum likelihood approach inherent in the joint model, which accommodates missingness under the missing-at-random assumption. We conducted a pattern-mixture model sensitivity analysis to assess robustness to missing-not-at-random mechanisms.

### Dynamic predictions

We computed individualized dynamic predictions of discontinuation probability at future time horizons (3, 6, and 12 months ahead), conditional on each participant’s observed longitudinal IBS-SSS history, using the methodology of Rizopoulos (2011) [19].

Predictive accuracy was evaluated using time-dependent area under the receiver operating characteristic curve (AUC) and expected Brier score via 10-fold cross-validation.

### Sample size

With 2,847 participants, 1,042 events, and six longitudinal measurements per participant (17,082 observations), the study exceeds recommended thresholds of 100 events and 500 subjects for stable joint model estimation [20].

## Results

### Participant Characteristics

Of 2,847 enrolled participants, 1,836 (64.5%) were female, mean age was 42.3 years (SD 13.7), and mean symptom duration was 7.8 years (SD 6.2). The IBS subtype distribution was: IBS-D 38.2%, IBS-C 27.5%, IBS-M 28.1%, and IBS-U 6.2%. Mean baseline IBS-SSS was 298.4 (SD 72.1), with 43.7% classified as severe. Clinically significant anxiety (HADS-A ≥8) was present in 41.2% and depression (HADS-D ≥8) in 23.6%. Social media use for IBS health information was reported by 52.8% of participants. Table 1 presents the full baseline characteristics stratified by treatment discontinuation status.

**Table 1.** Baseline Characteristics by Treatment Discontinuation Status (N = 2,847)

| Characteristic | Continued (n=1,805) | Discontinued | p-value |
| --- | --- | --- | --- |

|  |  | (n=1,042) |  |
| --- | --- | --- | --- |
| Age, mean (SD), years | 43.1 (13.9) | 40.8 (13.2) | 0.003 |
| Female sex, n (%) | 1,152 (63.8) | 684 (65.6) | 0.33 |
| **IBS subtype, n (%)** |  |  | **<0.001** |
| IBS-D | 648 (35.9) | 440 (42.2) |  |
| IBS-C | 512 (28.4) | 271 (26.0) |  |
| IBS-M | 528 (29.3) | 272 (26.1) |  |
| IBS-U | 117 (6.5) | 59 (5.7) |  |
| Baseline IBS-SSS, mean (SD) | 287.6 (69.3) | 317.1 (73.8) | <0.001 |
| Symptom duration, mean (SD), years | 7.5 (5.9) | 8.3 (6.7) | 0.02 |
| HADS-Anxiety $\geq 8$, n (%) | 684 (37.9) | 489 (46.9) | <0.001 |
| HADS-Depression $\geq 8$, n (%) | 387 (21.4) | 285 (27.4) | <0.001 |
| Low-FODMAP diet adherence, n (%) | 612 (33.9) | 298 (28.6) | 0.004 |
| Social media health info use, n (%) | 987 (54.7) | 516 (49.5) | 0.008 |
| Prior treatments, mean (SD) | 2.8 (1.9) | 3.4 (2.3) | <0.001 |

*IBS-SSS, Irritable Bowel Syndrome Severity Scoring System; HADS, Hospital Anxiety and Depression Scale; IBS-D, diarrhoea-predominant; IBS-C, constipation-predominant; IBS-M, mixed; IBS-U, unsubtyped; FODMAP, fermentable oligosaccharides, disaccharides, monosaccharides and polyols.*

### Longitudinal Submodel

The longitudinal submodel revealed a mean population-level decline in IBS-SSS of −8.7 points per month (95% CI: −10.2, −7.1; p < 0.001), corresponding to approximately a 52-point improvement over the first six months. However, the random effects variance components indicated substantial between-person heterogeneity: the random intercept variance was σ^2^(b□) = 4,218.3 (SD = 64.9) and the random slope variance was σ^2^(b□) = 12.4 (SD = 3.5), with a negative correlation between intercepts and slopes (r = −0.31, 95% CI: −0.38, −0.24), indicating that individuals with higher baseline severity tended to show steeper improvement trajectories. The residual variance was σ^2^ = 1,847.2. Significant fixed-effect covariates included IBS subtype (IBS-D showed 23.4 points higher severity than IBS-C at baseline, p < 0.001), concurrent anxiety (HADS-A ≥8 associated with 18.7 points higher IBS-SSS, p < 0.001), and low-FODMAP diet adherence (−15.3 points, p = 0.002). The time-by-anxiety interaction indicated that anxious participants showed slower improvement (β = 1.8 points/month attenuation, p = 0.014).

### Survival Submodel

The median time to treatment discontinuation was 16.8 months (95% CI: 15.2, 18.5). The Weibull shape parameter was κ = 1.23 (95% CI: 1.14, 1.33), indicating a modestly increasing hazard over time. In the survival submodel of the joint model, significant predictors of discontinuation included IBS-D subtype (HR = 1.41; 95% CI: 1.18, 1.69; p < 0.001), number of prior treatments (HR = 1.08 per additional treatment; 95% CI: 1.03, 1.13; p = 0.002), concurrent anxiety (HR = 1.28; 95% CI: 1.09, 1.50; p = 0.003), and depression (HR = 1.22; 95% CI: 1.01, 1.47; p = 0.038). Low-FODMAP diet adherence was protective (HR = 0.79; 95% CI: 0.67, 0.93; p = 0.005), as was social media use for IBS health information (HR = 0.82; 95% CI: 0.71, 0.95; p = 0.008). Table 2 presents the complete joint model results.

**Table 2.** Joint Longitudinal-Survival Model Parameter Estimates.

| Parameter | Estimate | 95% CI | p-value |
| --- | --- | --- | --- |
| **Longitudinal Submodel (IBS-SSS)** |  |  |  |
| Intercept ($\beta_0$) | 301.2 | 293.8, 308.6 | <0.001 |
| Time (months) ($\beta_1$) | −8.7 | −10.2, −7.1 | <0.001 |
| IBS-D (vs. IBS-C) | 23.4 | 14.1, 32.7 | <0.001 |
| Anxiety (HADS-A $\geq 8$) | 18.7 | 11.2, 26.2 | <0.001 |
| Low-FODMAP adherence | −15.3 | −25.0, −5.6 | 0.002 |
| Time × Anxiety interaction | 1.8 | 0.4, 3.2 | 0.014 |
| **Survival Submodel (Discontinuation)** |  |  |  |
|  | **HR** | **95% CI** |  |
| IBS-D subtype | 1.41 | 1.18, 1.69 | <0.001 |
| Prior treatments (per unit) | 1.08 | 1.03, 1.13 | 0.002 |
| Anxiety (HADS-A $\geq 8$) | 1.28 | 1.09, 1.50 | 0.003 |
| Depression (HADS-D $\geq 8$) | 1.22 | 1.01, 1.47 | 0.038 |
| Low-FODMAP adherence | 0.79 | 0.67, 0.93 | 0.005 |
| Social media health info use | 0.82 | 0.71, 0.95 | 0.008 |
| **Association Parameters** |  |  |  |
| $\alpha_{\square}$ (shared intercept) | 0.0034 | 0.0021, 0.0047 | <0.001 |
| $\alpha_{\square}$ (shared slope) | -0.187 | -0.264, -0.110 | <0.001 |

*HR, hazard ratio; CI, confidence interval; IBS-SSS, Irritable Bowel Syndrome Severity Scoring System; IBS-D, diarrhoea-predominant; IBS-C, constipation-predominant; HADS, Hospital Anxiety and Depression Scale; FODMAP, fermentable oligosaccharides, disaccharides, monosaccharides and polyols.*

**Table 3.**
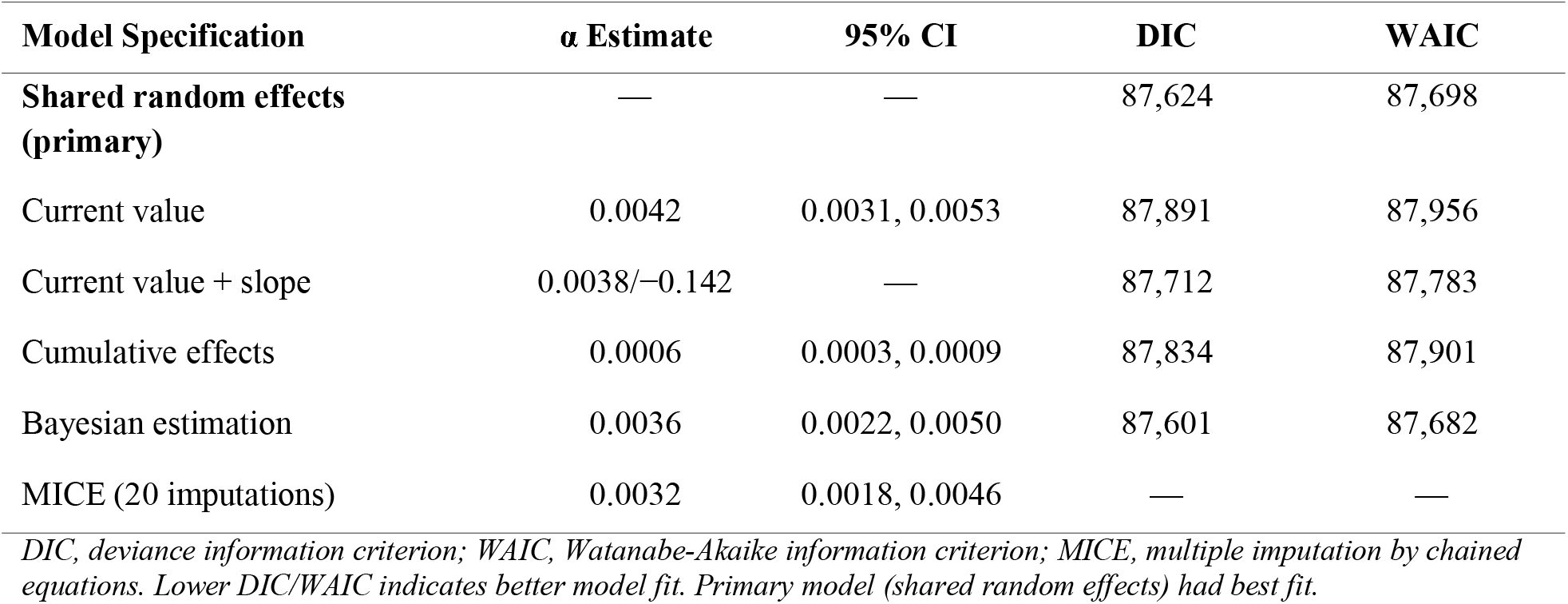
Comparison of Joint Model Association Structures and Sensitivity Analyses.

### Association Parameters and Joint Model Inference

The association parameters from the joint model provided the central finding of this study. The shared random intercept association parameter was α□ = 0.0034 (95% CI: 0.0021, 0.0047; p < 0.001), indicating that each 10-point increase in the individual-specific baseline IBS-SSS (above the population mean) was associated with a 3.5% increase in the hazard of treatment discontinuation (exp(0.0034 × 10) = 1.035). The shared random slope association parameter was α□ = −0.187 (95% CI: −0.264, −0.110; p < 0.001), demonstrating that individuals with steeper negative slopes (i.e., faster symptom improvement) had substantially lower discontinuation hazards. A one-unit increase in the individual-specific monthly improvement rate (more negative slope) was associated with a 17.1% reduction in the hazard of discontinuation (exp(−0.187) = 0.829).

Comparing the joint model to separate models, the joint model produced notably different estimates for the longitudinal trajectory. The separate LME model estimated the population slope as −7.3 points/month (95% CI: −8.6, −6.0), while the joint model estimated −8.7 points/month.

This 19% difference reflects the bias introduced by informative dropout: participants who discontinued (and thus contributed fewer observations) tended to have worse trajectories, and ignoring this non-random missingness attenuated the estimated treatment effect. The log-likelihood comparison confirmed superior fit of the joint model (log-L = −43,812.4) over the sum of separate models (log-L = −44,287.1; Δlog-L = 474.7; p < 0.001 by likelihood ratio test).

### Sensitivity Analyses and Model Diagnostics

The shared random effects parameterization demonstrated the best fit across all metrics (DIC = 87,624; WAIC = 87,698), outperforming the current value (DIC = 87,891), current value plus slope (DIC = 87,712), and cumulative effects (DIC = 87,834) specifications. Bayesian estimation with weakly informative priors (Normal(0, 100) for fixed effects, half-Cauchy(0, 2.5) for variance components) yielded substantively identical results (α□ = 0.0036, 95% CrI: 0.0022, 0.0050), confirming robustness to estimation method. The MICE sensitivity analysis produced consistent association estimates (α□ = 0.0032, 95% CI: 0.0018, 0.0046). The pattern-mixture model incorporating δ = 10 and δ = 20 point departures from MAR yielded qualitatively unchanged conclusions, with α□ ranging from 0.0029 to 0.0039.

### Dynamic Prediction Performance

Dynamic predictions demonstrated strong discriminative ability. Table 4 presents the time-dependent AUC and Brier scores for predicting discontinuation at 3-, 6-, and 12-month horizons, conditional on varying amounts of longitudinal data.

**Table 4.** Dynamic Prediction Performance: Time-Dependent AUC and Brier Scores.

| Landmark Time | Horizon | AUC (95% CI) | Brier Score | Brier (Separate) |
| --- | --- | --- | --- | --- |
| 3 months | +3 months | 0.72 (0.68, 0.76) | 0.187 | 0.214 |
| 3 months | +6 months | 0.70 (0.66, 0.74) | 0.198 | 0.228 |
| 6 months | +3 months | 0.76 (0.72, 0.80) | 0.172 | 0.201 |
| 6 months | +6 months | 0.74 (0.70, 0.78) | 0.181 | 0.209 |
| 12 months | +6 months | 0.79 (0.75, 0.83) | 0.158 | 0.189 |
| 12 months | +12 months | 0.77 (0.73, 0.81) | 0.164 | 0.196 |

*AUC, area under the receiver operating characteristic curve; CI, confidence interval. AUC and Brier scores estimated via 10-fold cross-validation. Lower Brier scores indicate better calibration. Separate model refers to prediction from independent longitudinal and survival models.*

**Table 5.** Association Parameter Estimates by Subgroup.

| Subgroup | n (events) | $\alpha_{\square}$ (intercept) | $\alpha_{\square}$ (slope) | p-interaction |
| --- | --- | --- | --- | --- |
| Overall | 2,847 (1,042) | 0.0034 | −0.187 | — |
| IBS-D | 1,088 (440) | 0.0041 | −0.224 | 0.032 |
| IBS-C | 783 (271) | 0.0028 | −0.156 | ref |
| IBS-M | 800 (272) | 0.0033 | −0.178 | 0.41 |
| Anxiety (HADS-A $\geq 8$) | 1,173 (489) | 0.0047 | −0.231 | 0.009 |
| No anxiety | 1,674 (553) | 0.0024 | −0.148 | ref |
| Social media user | 1,503 (516) | 0.0029 | −0.162 | 0.048 |
| Non-social media user | 1,344 (526) | 0.0040 | −0.218 | ref |

*p-interaction tests whether the association parameter differs significantly from the reference subgroup. IBS-D, diarrhoea-predominant; IBS-C, constipation-predominant; IBS-M, mixed; HADS, Hospital Anxiety and Depression Scale.*

Predictive accuracy improved with additional longitudinal data: the 12-month landmark AUC for 6-month-ahead prediction was 0.79 (95% CI: 0.75, 0.83), compared to 0.72 (95% CI: 0.68, 0.76) at the 3-month landmark. Notably, the joint model consistently outperformed separate models in calibration (lower Brier scores), with the largest improvements observed at later landmarks where the informative dropout mechanism was most pronounced.

### Subgroup and Heterogeneity Analyses

Subgroup analyses revealed significant effect modification. The symptom trajectory– discontinuation association was strongest among IBS-D patients (α□ = −0.224 vs. −0.156 for IBS-C; p-interaction = 0.032) and among participants with concurrent anxiety (α□ = −0.231 vs. −0.148; p-interaction = 0.009). Notably, social media users for IBS health information showed a weaker trajectory–discontinuation link (α□ = −0.162 vs. −0.218; p-interaction = 0.048), suggesting that access to online peer support and health information may partially buffer the impact of inadequate symptom improvement on treatment persistence decisions.

## Discussion

This study applied joint longitudinal-survival modelling to a large prospective Canadian IBS cohort, demonstrating that patient-reported symptom trajectories and treatment discontinuation are dynamically linked through shared individual-level latent processes. The shared random effects association parameters reveal that both the level and rate of change in symptom severity independently predict discontinuation hazards, with meaningful clinical implications for treatment monitoring and personalized care.

Our finding that higher individual-specific baseline severity predicts earlier discontinuation (α□ = 0.0034) extends prior cross-sectional evidence by showing that this association operates through latent individual differences rather than merely observed baseline scores. This distinction is important: the random intercept captures unmeasured patient characteristics (e.g., pain sensitivity, illness cognition, treatment expectations) that jointly determine initial severity and persistence behaviour. The stronger slope association (α□ = −0.187), indicating that each unit increase in monthly improvement rate reduces discontinuation hazard by 17.1%, provides the first quantitative evidence that real-time symptom dynamics—not just static baseline features—drive treatment decisions in IBS.

The social media health information finding merits particular attention. Participants who used social media for IBS-related health information showed both lower discontinuation hazards (HR = 0.82) and a weaker trajectory–discontinuation association (α□ = −0.162 vs. −0.218). This aligns with the comprehensive social media discourse analysis by Shankar and Yip (2025), who identified social support (14.2% of discourse) and awareness-raising (11.5%) as major thematic domains in IBS-related online discussions, alongside more medically oriented topics such as dietary management (15.1%) and treatment approaches (12.2%) [10]. Their finding that the majority of social media discourse was neutral (45.9%) or positive (35.4%) in sentiment, reflecting information-sharing and support-seeking behaviour rather than negative emotional expression, supports our observation that social media engagement may foster treatment persistence through peer validation, shared coping strategies, and normalized illness experience.

The comparison between joint and separate models underscores the methodological importance of accounting for informative dropout. The 19% underestimation of the population-level improvement slope by the separate LME model represents clinically meaningful bias that could lead to pessimistic conclusions about treatment effectiveness. This bias arises because patients with inadequate improvement are selectively lost to follow-up through discontinuation, truncating the poorer-performing end of the trajectory distribution. Joint modelling corrects this selection effect by simultaneously estimating the dropout mechanism, yielding less biased treatment effect estimates.

The dynamic prediction framework offers practical clinical utility. At the 12-month landmark, the joint model achieved an AUC of 0.79 for predicting 6-month-ahead discontinuation, outperforming separate models (AUC improvement of 0.04–0.06 depending on horizon). This discriminative ability, combined with the individualized nature of dynamic predictions that update as new longitudinal data accumulate, suggests a viable pathway toward clinical decision support tools that flag patients at elevated discontinuation risk based on their evolving symptom trajectory.

The subgroup heterogeneity findings have direct clinical implications. The stronger trajectory– discontinuation coupling in IBS-D patients and those with concurrent anxiety identifies populations requiring intensified monitoring and potentially earlier intervention adjustment.

These findings echo the prominence of mental health themes in patient online discourse [10], where anxiety and chronic comorbidities feature as distinct discussion topics, suggesting that patients themselves recognize the interplay between psychological wellbeing and IBS management.

## Strengths and Limitations

Key strengths include the large sample size (n = 2,847 with 1,042 events), multicentre design across three provinces, validated outcome measures, comprehensive sensitivity analyses (alternative association structures, Bayesian estimation, MICE, pattern-mixture models), and the dynamic prediction framework with cross-validated performance assessment. The 24-month follow-up with six measurement occasions provided sufficient longitudinal resolution to estimate individual trajectories reliably.

Several limitations warrant consideration. First, treatment discontinuation was defined as cessation for ≥30 consecutive days, which may not capture all forms of non-adherence (e.g., dose reduction, treatment holidays). Second, while the multicentre design enhances generalizability within Canada, the cohort was drawn from tertiary gastroenterology centres, potentially representing more severe cases than the general IBS population. Third, the linear trajectory assumption may not capture non-linear recovery patterns; future work could extend to spline-based or piecewise trajectories within the joint framework. Fourth, social media use was assessed as a binary self-report variable, precluding more granular analysis of engagement intensity, content exposure, or the specific mechanisms through which online health information seeking influences treatment persistence. Fifth, the observational design limits causal inference regarding the social media–persistence association.

### Implications for Practice and Research

These findings support three actionable recommendations. First, clinicians managing IBS should implement routine symptom trajectory monitoring using validated instruments such as the IBS-SSS at regular intervals, as the rate of early improvement is a stronger predictor of long-term persistence than baseline severity alone. Second, targeted interventions for patients at high discontinuation risk—identified by IBS-D subtype, concurrent anxiety, high baseline severity, and slow early improvement—should include proactive follow-up, expectation management, and potentially earlier treatment modification. Third, the protective association of social media health information use suggests that healthcare systems should consider integrating curated online peer support resources and patient education platforms into IBS management pathways, consistent with the rich information-sharing ecosystem documented in social media discourse analyses [10].

## Conclusion

Joint longitudinal-survival modelling reveals that IBS symptom trajectories and treatment discontinuation are dynamically linked through individual-level latent processes that cannot be captured by separate analyses. Higher baseline severity and slower improvement trajectories significantly predict earlier treatment discontinuation, with the association moderated by IBS subtype, anxiety comorbidity, and social media health information engagement. These findings advance understanding of treatment persistence in functional gastrointestinal disorders and provide a statistical and clinical framework for personalized monitoring that leverages real-time patient-reported outcome data to identify at-risk individuals and inform timely intervention adaptation.

## Data Availability

All data produced in the present work are contained in the manuscript

